# Estimating Hepatitis C Virus Prevalence in US States and the District of Columbia, 2017–2020

**DOI:** 10.64898/2026.09.29.26364309

**Authors:** Heather Bradley, Ya-Hui Yu, Lanxin Li, Shashi N. Kapadia, Patrick S. Sullivan, Eric W. Hall

**Author notes:** Authors contributed equally to this work (Heather Bradley, Ya-Hui Yu). Correspondence: Heather Bradley.

## Abstract

**Background:** State-level estimates of hepatitis C virus (HCV) prevalence are needed to guide resource allocation and to provide a baseline against which to measure progress toward elimination goals. Population prevalence is not directly observable, requiring estimation from indirect data sources.

**Methods:** We adapted a Bayesian spatial integrated abundance model to estimate state-level HCV prevalence during 2017–2020 across 48 states and the District of Columbia. The model integrated six HCV-related outcomes: acute and chronic surveillance cases, HCV-related deaths, observations of HCV in [MarketScan] administrative claims data, diagnoses of HCV in Medicaid recipients, and treatment with direct-acting antivirals in Medicaid recipients. Estimates were anchored to a national prevalence estimate, and the model accounted for data source–specific selection, heterogeneity in HCV risk factors across states, and geospatial correlation.

**Results:** Estimated average prevalence was 1.32% (95% credible interval [CrI]: 0.94%–1.80%), corresponding to 3.33 million (95% CrI: 2.37–4.65 million) adults with HCV infection across 48 states and DC. State estimates ranged from 0.74% in North Dakota to 2.22% in Oklahoma (median state-specific prevalence, 1.27%), with higher prevalence concentrated in South Central states, Appalachia, and the West, and the lower prevalence in the upper Midwest, Southeast, and New England. Ten states accounted for 56% of estimated infections. Estimates were stable in sensitivity analyses (most differences <1 percentage point).

**Conclusions:** These estimates quantify state-level HCV burden prior to the 2021 federal expansion of HCV surveillance funding, providing a baseline for monitoring elimination progress. Geographic variation indicates that resource needs will differ by jurisdiction.

## Introduction

Hepatitis C virus (HCV) is the most common bloodborne infectious disease in the U.S. and causes substantial liver-related morbidity and mortality. A recent study estimated up to 4 million people in the U.S. had HCV in 2017 – 2020,^1^ and more than 11,000 people die annually from HCV-related liver disease^2^. However, HCV-related mortality rates declined by more than 50% during the last decade as direct-acting antiviral (DAA) curative medications were introduced^3^. Given this success and following the lead of the World Health Organization, the U.S. released a plan in 2020 to eliminate HCV by 2030^4^.

Unfortunately, the U.S. is not likely to reach this goal by 2030. Before 2011, HCV infections were concentrated among persons born from 1945 – 1965, most of whom acquired infection before the blood supply was routinely screened^5^. Acute HCV infections began increasing again in 2011, and most were reported among people <40 years old. Injection drug use is now the most common risk factor for acute HCV infection^6,7^. This increase in new HCV infections is a devastating consequence of the U.S. opioid crisis^8^. Harm reduction and treatment services have not kept pace with population needs^9,10^.

Although HCV-related mortality has declined overall, important inequities persist: males, American Indian/Alaska Native persons, Black persons, and 55 – 74-year-olds have the highest mortality rates^2^.

There have been — as yet unsuccessful — efforts to fund a national HCV elimination plan; the Cure Hepatitis C Act, is bipartisan legislation introduced in 2025, but not yet passed^11^. Although resources for prevention and treatment of HCV infections are critical, they will not be sufficient to eliminate HCV. As interventions are implemented, HCV burden must be monitored at sub-national levels and in high-risk groups. HCV surveillance is chronically under-resourced. Before 2021, only 14 U.S. jurisdictions (i.e., states and large cities) received federal funding for hepatitis surveillance. One-third of jurisdictions did not have even one full-time employee committed to hepatitis surveillance^12^. In 2021, CDC expanded hepatitis surveillance funding to 59 jurisdictions, allowing states and cities to begin expanding surveillance capacity and working toward elimination goals^13^.

Because of surveillance limitations and the large proportion of infections that are undiagnosed^14^, most data on HCV prevalence are derived from models^15^. The most recent state-level modeled estimates represent 2013 – 2016^16^. In this manuscript, benchmarking to the recently published 2017 – 2020 national estimate, we estimate state-level prevalence during this period, which directly precedes expanded surveillance funding and introduction of federal and local elimination strategies. We aim to provide HCV prevalence estimates that are comparable across U.S. states to help guide federal resource allocation and to provide states with a baseline against which they can compare progress toward elimination goals. For the purposes of this study, we refer to HCV prevalence as the percent of the population with current HCV infection that has neither been cleared nor cured, i.e., viremic prevalence.

## Methods

To estimate comparable state-level HCV prevalence, we adapted a Bayesian spatiotemporal integrated abundance model^17,18^, a hierarchical modelling framework for estimating hidden populations from partially observed aggregate counts. This approach has recently been applied to estimate county-level prevalence of opioid misuse^17,19,20^ and is well suited for HCV because HCV-related data sources capture incomplete and distinct parts of the underlying population with current infection.

Our model incorporated multiple HCV-related outcomes, anchored to a national HCV prevalence estimate, with separate state-level covariate sets modeling data source–specific selection and heterogeneity in HCV risk. The model also integrated a spatial component that accounted for correlation across neighboring jurisdictions defined using adjacency among geographically connected jurisdictions. Analyses were therefore restricted to the 48 contiguous states and the District of Columbia.

### Data sources

We identified six state-level HCV-related outcomes that represented partial counts of the population with current HCV infection during 2017-2020. The first two outcomes were annual counts of acute and chronic HCV cases, which met surveillance case definitions created by Centers for Disease Control and Prevention (CDC)/Council of State and Territorial Epidemiologists and were reported through state and local surveillance systems to CDC^21^. Acute HCV surveillance cases^22,23^ represent newly identified infections with clinical or laboratory evidence of recent infection, whereas chronic HCV surveillance cases^24,25^ are identified by laboratory evidence of current or past HCV infection without evidence of acute disease. The third outcome was cumulative HCV-related deaths reported to the National Vital Statistics System^26^. Deaths were defined by having ICD-10 codes for acute viral hepatitis C (B17.1) or chronic viral hepatitis C (B18.2) as an underlying or contributing cause of death.

The other three HCV-related outcomes were identified through large administrative databases. The fourth outcome was a cumulative count of adults with evidence of HCV infection in the Merative™ MarketScan® Commercial and Medicare Databases, a convenience sample of deidentified, person-level U.S. health care claims data from primarily privately insured individuals. The Medicare component in MarketScan includes Medicare-eligible retirees with employer-sponsored supplemental coverage. Evidence of HCV infection was defined by the presence of an HCV diagnostic code or an National Drug Code for DAA treatment during the study period (see codes in eAppendix S1). The fifth outcome was a cumulative count of Medicaid recipients with a new HCV diagnosis, as defined in a prior study^27^. We used aggregated state-level counts from this published study, therefore this outcome was limited to the original study period of January 1, 2018 through June 30, 2019. The last outcome was the cumulative number of Medicaid recipients who were treated with a DAA between 2017-2020, as reported in a prior study^28^. Additional details on outcome definitions and the complete lists of diagnosis and medication codes used for these outcomes are provided in eAppendix S1.

We used a recently published national estimate of HCV period prevalence for 2017–2020^1^ (1.6%; 95% CI: 0.9%–2.2%) as a benchmark to inform the model: This modeled estimate was based on HCV RNA prevalence in National Health and Nutrition Examination Survey (NHANES) data, with additional adjustment for the underrepresentation of people who inject drugs and other populations excluded from the NHANES sampling frame. The adult population for each state was calculated as the average population aged ≥18 years during 2017–2020 using CDC WONDER population estimates^29^.

Two types of state-level covariates were incorporated into the model. First, outcome-specific covariates were included to account for factors associated with the state-level proportion of adults with HCV infection that was likely to be captured by each observed outcome. For acute and chronic HCV surveillance cases, we accounted for surveillance capacity and reporting completeness by including data on the average surveillance funding received by state^30^ during 2017-2020, and the number of years with HCV surveillance data reported to CDC^21^. For HCV-related deaths, we included measures of medicolegal death investigation systems for each state^31^ to account for potential variation in cause-of-death ascertainment across death investigation systems.

For the HCV infections in MarketScan, we used the ratio of MarketScan enrollees to the adult population in each state as a proxy for the proportion of adults in a state who are captured by MarketScan. Because MarketScan is a convenience sample, we also included demographic and socioeconomic covariates^32^ to account for potential state-level differences in the characteristics of insured versus uninsured adults and the extent to which underlying HCV infections were likely to be observed through diagnosis or DAA treatment claims in the database^33^. The Medicaid-related outcomes included age distribution and covariates representing Medicaid enrollment as a percentage of the adult population, Medicaid expansion status, and DAA treatment policies^28^. These covariates may affect whether adults with HCV are enrolled in Medicaid, diagnosed with HCV, or receive DAA treatment.

The second type of state-level covariates consisted of prevalence covariates, which were included to characterize state-level differences in underlying risk factors for HCV infection. We included sociodemographic composition^32^, indicators of structural vulnerability (such as unsheltered homelessness^34^ and adult incarceration^35^), and risk factors related to injection drug use–associated harms^36^, including drug overdose deaths^37^ and treatment admissions reporting injection drug use^38^. The complete list of covariates is provided in Table 1; definitions and data sources are described in eAppendix S1.

**Table 1.**
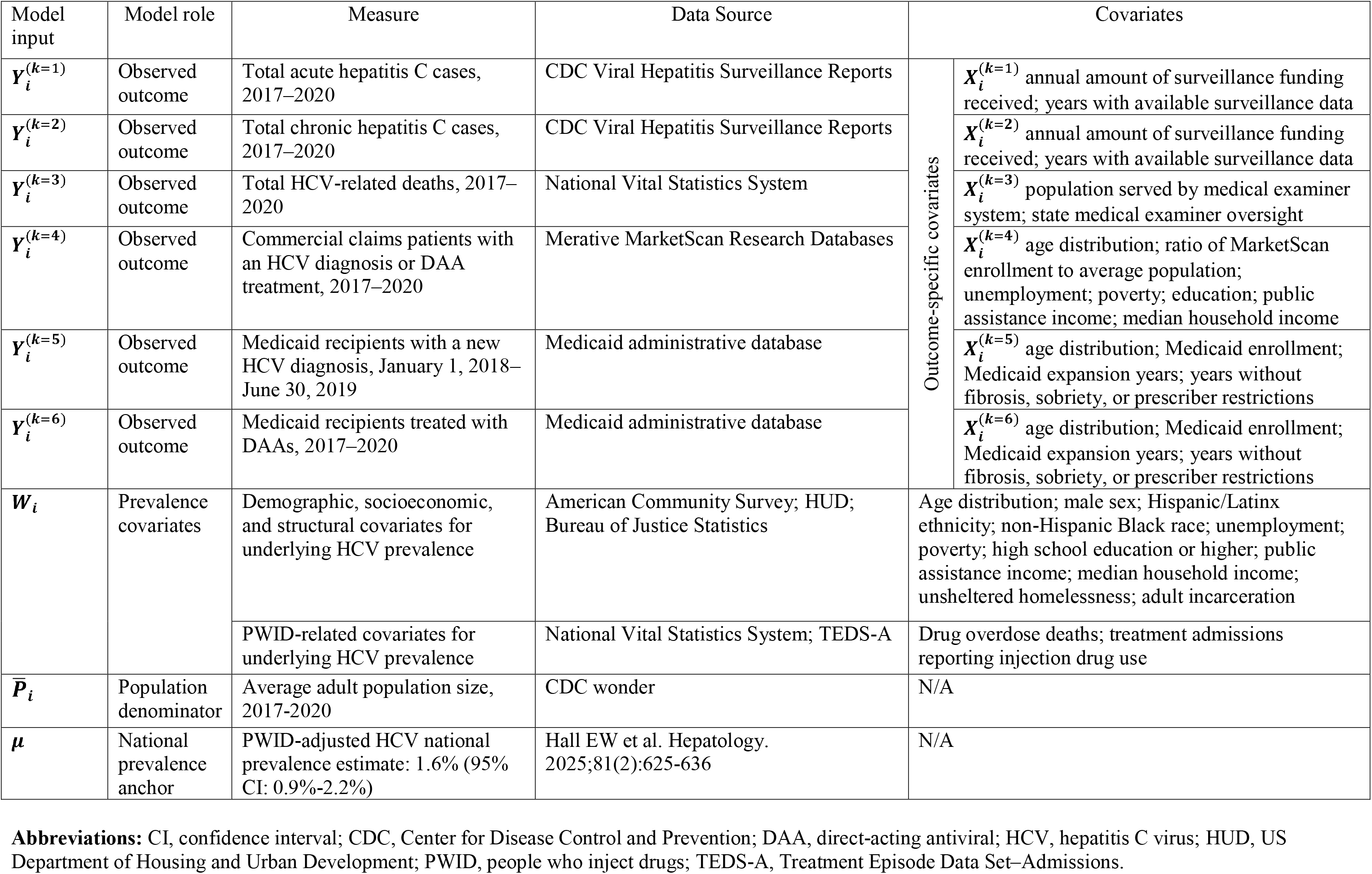
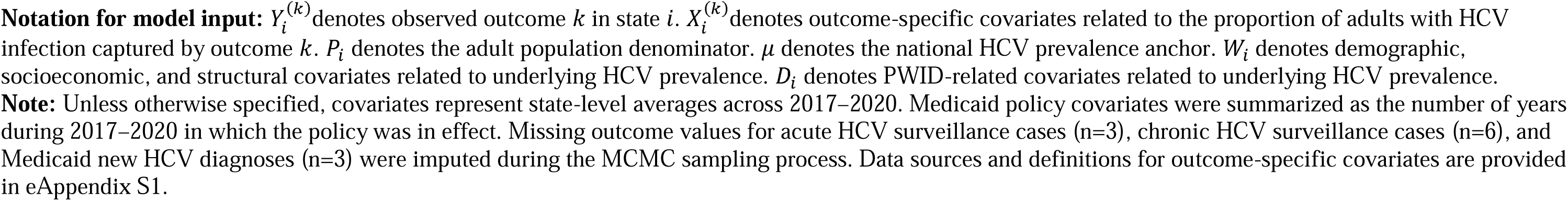
Overview of model inputs used in the Bayesian spatial integrated abundance model, 48 states and the District of Columbia, 2017–2020.

| Model input | Model role | Measure | Data Source | Covariates |  |
| --- | --- | --- | --- | --- | --- |
| $Y_i^{(k=1)}$ | Observed outcome | Total acute hepatitis C cases, 2017–2020 | CDC Viral Hepatitis Surveillance Reports | Outcome-specific covariates | $X_i^{(k=1)}$ annual amount of surveillance funding received; years with available surveillance data |
| $Y_i^{(k=2)}$ | Observed outcome | Total chronic hepatitis C cases, 2017–2020 | CDC Viral Hepatitis Surveillance Reports | | $X_i^{(k=2)}$ annual amount of surveillance funding received; years with available surveillance data |
| $Y_i^{(k=3)}$ | Observed outcome | Total HCV-related deaths, 2017–2020 | National Vital Statistics System | | $X_i^{(k=3)}$ population served by medical examiner system; state medical examiner oversight |
| $Y_i^{(k=4)}$ | Observed outcome | Commercial claims patients with an HCV diagnosis or DAA treatment, 2017–2020 | Merative MarketScan Research Databases | | $X_i^{(k=4)}$ age distribution; ratio of MarketScan enrollment to average population; unemployment; poverty; education; public assistance income; median household income |
| $Y_i^{(k=5)}$ | Observed outcome | Medicaid recipients with a new HCV diagnosis, January 1, 2018–June 30, 2019 | Medicaid administrative database | | $X_i^{(k=5)}$ age distribution; Medicaid enrollment; Medicaid expansion years; years without fibrosis, sobriety, or prescriber restrictions |
| $Y_i^{(k=6)}$ | Observed outcome | Medicaid recipients treated with DAAs, 2017–2020 | Medicaid administrative database | | $X_i^{(k=6)}$ age distribution; Medicaid enrollment; Medicaid expansion years; years without fibrosis, sobriety, or prescriber restrictions |
| $W_i$ | Prevalence covariates | Demographic, socioeconomic, and structural covariates for underlying HCV prevalence | American Community Survey; HUD; Bureau of Justice Statistics | Age distribution; male sex; Hispanic/Latinx ethnicity; non-Hispanic Black race; unemployment; poverty; high school education or higher; public assistance income; median household income; unsheltered homelessness; adult incarceration | |
|  |  | PWID-related covariates for underlying HCV prevalence | National Vital Statistics System; TEDS-A | Drug overdose deaths; treatment admissions reporting injection drug use |  |
| $\bar{P}_i$ | Population denominator | Average adult population size, 2017–2020 | CDC wonder | N/A | |
| $\mu$ | National prevalence anchor | PWID-adjusted HCV national prevalence estimate: 1.6% (95% CI: 0.9%–2.2%) | Hall EW et al. Hepatology. 2025;81(2):625–636 | N/A | |

**Abbreviations:** CI, confidence interval; CDC, Center for Disease Control and Prevention; DAA, direct-acting antiviral; HCV, hepatitis C virus; HUD, US Department of Housing and Urban Development; PWID, people who inject drugs; TEDS-A, Treatment Episode Data Set–Admissions.
**Notation for model input:** $Y_i^{(k)}$ denotes observed outcome $k$ in state $i$ . $X_i^{(k)}$ denotes outcome-specific covariates related to the proportion of adults with HCV infection captured by outcome $k$ . $P_i$ denotes the adult population denominator. $\mu$ denotes the national HCV prevalence anchor. $W_i$ denotes demographic, socioeconomic, and structural covariates related to underlying HCV prevalence. $D_i$ denotes PWID-related covariates related to underlying HCV prevalence.
**Note:** Unless otherwise specified, covariates represent state-level averages across 2017–2020. Medicaid policy covariates were summarized as the number of years during 2017–2020 in which the policy was in effect. Missing outcome values for acute HCV surveillance cases (n=3), chronic HCV surveillance cases (n=6), and Medicaid new HCV diagnoses (n=3) were imputed during the MCMC sampling process. Data sources and definitions for outcome-specific covariates are provided in eAppendix S1.

### Statistical analysis

We used a Bayesian spatial integrated abundance model, described briefly here, with input variables shown in Table 1, and additional model details are provided in eAppendix S2. We incorporated six HCV-related outcomes (*k*) in the model as described above. Counts were aggregated within each state (*i*) over the 2017–2020 study period (*t*) because the analysis estimated period prevalence rather than year-specific prevalence.

For each outcome (*k*), let 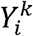 denote the total observed count in state i during the study period ***t***. We assume

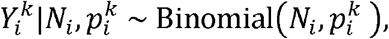

where *N*_*i*_ refers to the latent number of people with HCV infection in state *i* during ***t***, and 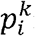 represents the rate of experiencing the outcome *k* in the population with HCV infection in state *i* during ***t***. The rate 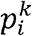 is modelled by

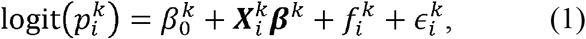

where 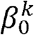 is the intercept, 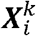 is the covariate vector related to state *i, **β***^*k*^ is the coefficient vector corresponding to 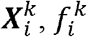 refers to the spatial random effect, and 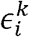 is the error term. To allow for potential correlation across outcomes, the error terms 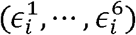 for each state *i* are assumed to be from a multivariate normal distribution with mean zero^20^. Further, let 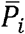 denote the averaged total population size in state *i* across the same time interval ***t***, we introduce

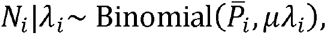

where *µ* refers to the national prevalence of HCV during ***t***, and *λi* is the prevalence ratio of HCV infection in state *i* compared with *µ*. Hence, the state-level HCV prevalence during *t* is *µ λ*_*i*_. The prevalence ratio *λ*_*i*_ is modelled by

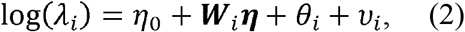

where *η*_0_ is the intercept, ***W***_*i*_ is the covariate vector related to state *i*, ***η*** is the coefficient vector, *θ*_*i*_ is the spatial random effect, and 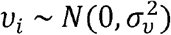 accounts for the unexplained variance. The spatial random effects 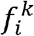 and *θ*_*i*_ were assumed to follow the intrinsic conditional autoregressive (ICAR) model^39^. Spatial relationships among the 48 states and the District of Columbia were defined using adjacency matrix based on shared geographic borders.

Using weakly informative prior settings for parameters in models (1) and (2), the full joint posterior distribution 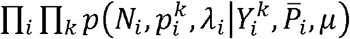 was simulated by a Markov chain Monte Carlo (MCMC) algorithm using the *R* package *NIMBLE*^40^. Ten MCMC chains were set up with different random initialisations. For each chain, the total number of iterations was set to 100,000. The first half of these draws were discarded as burn-in, and the remaining draws were thinned by 5 before inference. Convergence was checked using trace plots and the modified Gelman–Rubin diagnostic statistics^41^. Posterior means were used as point estimates, and 95% credible intervals (Crl) were calculated as the 2.5th and 97.5th percentiles of the posterior draws.

We conducted two sets of sensitivity analyses for comparison with the outcomes of the main analysis which used all six HCV-related outcomes. First, we refit the model with excluding surveillance data (acute and chronic HCV cases). Because the study period preceded expanded federal viral hepatitis surveillance funding, case-based surveillance data may have varied across states in completeness and comparability due to differences in surveillance infrastructure and capacity. We therefore used this analysis to evaluate estimates derived only from non-surveillance outcomes. Second, we conducted leave-one-outcome-out analyses, excluding each of the six outcomes in turn, to assess the impact of each individual data source on state-level prevalence estimates.

For states with missing annual acute or chronic HCV surveillance counts, we first imputed missing values as the jurisdiction-specific mean across available years. For Medicaid DAA treatment receipt, yearly counts fewer than 10 prescriptions in a state were suppressed (n=1) in the aggregated state-level counts obtained through a data sharing agreement; we assigned this suppressed value a count of 5, the midpoint of the possible range. Medicaid new HCV diagnosis values were unavailable for three states because those states were excluded from the source study due to missing key variables. After these data-processing steps, remaining missing outcome values for acute HCV surveillance cases (n=3), chronic HCV surveillance cases (n=6), and Medicaid new HCV diagnoses (n=3) were treated as missing and imputed during the MCMC sampling process.

Example R code for the analyses is provided in a GitHub repository (anonymized for peer review).

## Results

The estimated period prevalence of HCV infection among US adults varied across states during 2017–2020. The estimated average prevalence across the 48 states and District of Columbia was 1.32% (95% CrI: 0.94%–1.80%) which is equivalent to an estimated 3.33 million (95% CrI: 2.37–4.65 million) adults with HCV infection during 2017-2020. Estimated prevalences ranged from 0.73% in North Dakota to 2.22% in Oklahoma, with a median state-specific estimate of 1.27% (Missouri).

Estimated prevalence was highest in several South Central states, Appalachia, and some states in the West Census Region, and lowest in portions of the upper Midwest, Southeastern states, and New England (Table 2, Figure 1). Nine of 11 included states in the West Census Region had prevalence estimates above the national median. In contrast, only 3 (Indiana, Michigan, and Ohio) of 12 states in the Midwest Region and 2 (New York, Rhode Island) of 9 states in the Northeast region had prevalence estimates above the national median. The largest number of estimated HCV infections was in California (n=441,186, 95% CrI: 265,134–641,503), and five of the ten states with the largest numbers of estimated infections were located in the South Census Region. Overall, these ten states (CA, TX, FL, NY, OH, PA, MI, TN, GA, NC) accounted for 56% of all estimated HCV infections in the country.

**Table 2.**
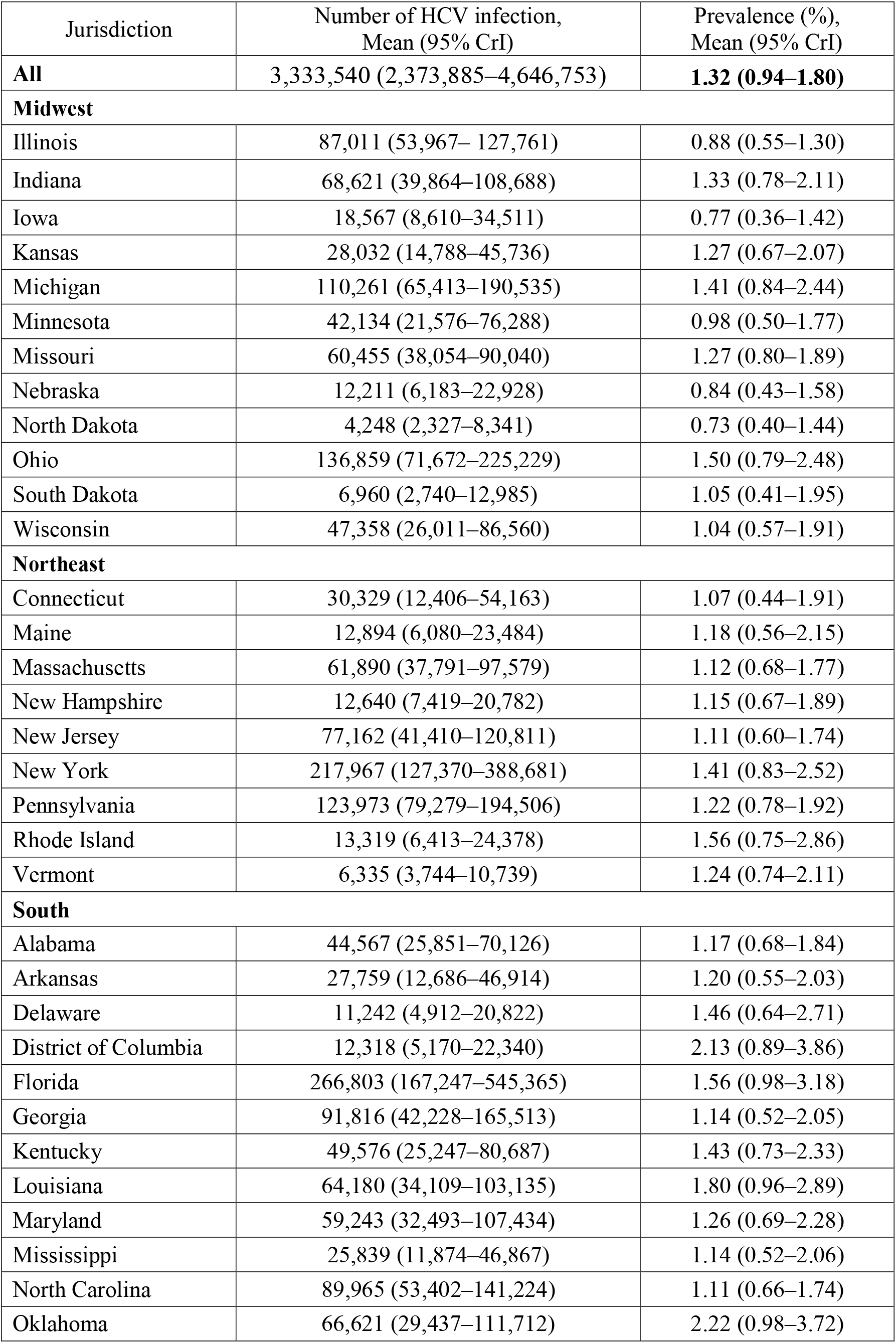
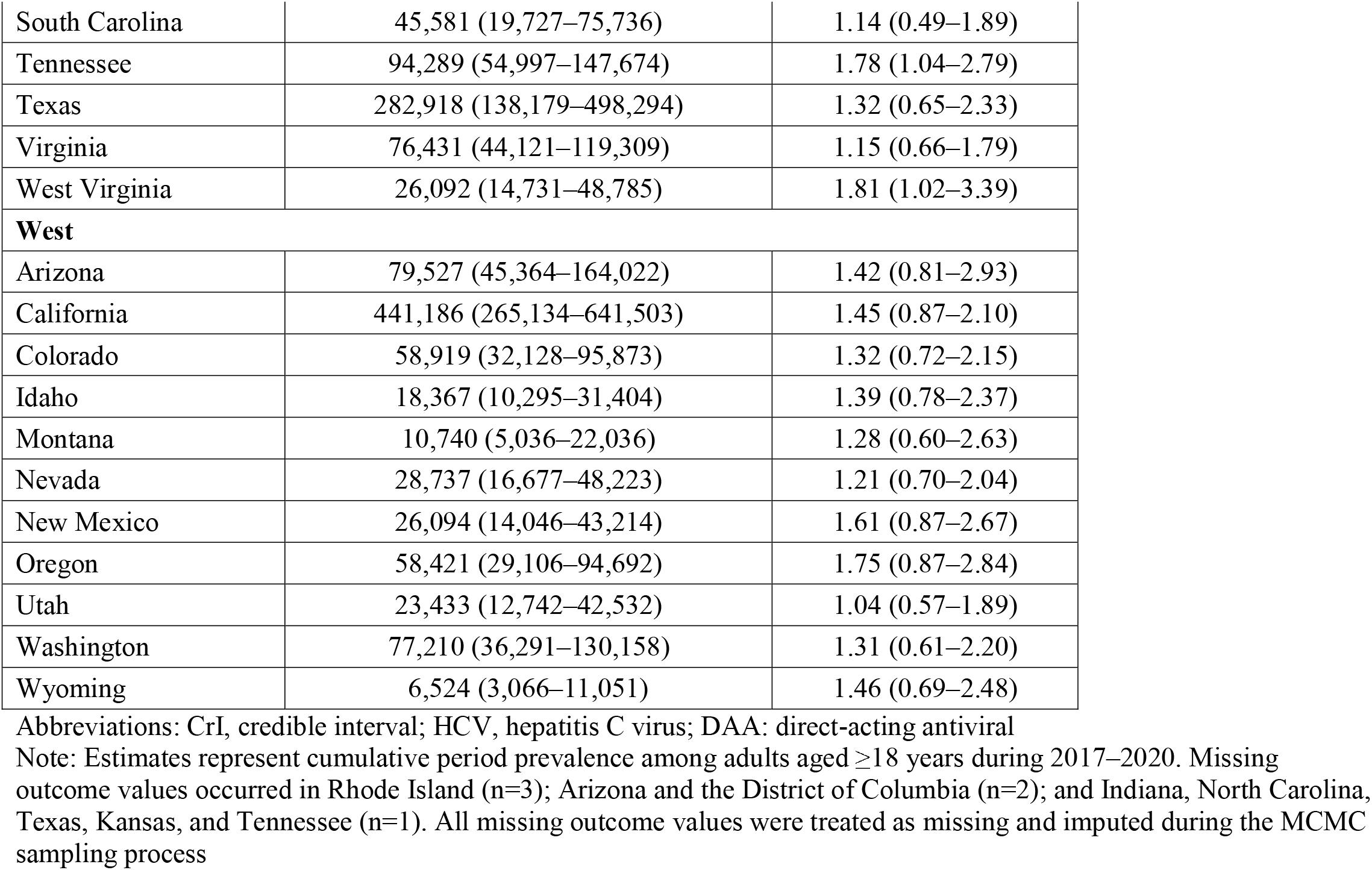
Estimated HCV prevalence (per 100 population) and number of adults with HCV infection by jurisdiction, 48 states and the District of Columbia, 2017–2020.

| Jurisdiction | Number of HCV infection,<br>Mean (95% CrI) | Prevalence (%),<br>Mean (95% CrI) |
| --- | --- | --- |
| **All** | 3,333,540 (2,373,885–4,646,753) | **1.32 (0.94–1.80)** |
| **Midwest** |  |  |
| Illinois | 87,011 (53,967–127,761) | 0.88 (0.55–1.30) |
| Indiana | 68,621 (39,864–108,688) | 1.33 (0.78–2.11) |
| Iowa | 18,567 (8,610–34,511) | 0.77 (0.36–1.42) |
| Kansas | 28,032 (14,788–45,736) | 1.27 (0.67–2.07) |
| Michigan | 110,261 (65,413–190,535) | 1.41 (0.84–2.44) |
| Minnesota | 42,134 (21,576–76,288) | 0.98 (0.50–1.77) |
| Missouri | 60,455 (38,054–90,040) | 1.27 (0.80–1.89) |
| Nebraska | 12,211 (6,183–22,928) | 0.84 (0.43–1.58) |
| North Dakota | 4,248 (2,327–8,341) | 0.73 (0.40–1.44) |
| Ohio | 136,859 (71,672–225,229) | 1.50 (0.79–2.48) |
| South Dakota | 6,960 (2,740–12,985) | 1.05 (0.41–1.95) |
| Wisconsin | 47,358 (26,011–86,560) | 1.04 (0.57–1.91) |
| **Northeast** |  |  |
| Connecticut | 30,329 (12,406–54,163) | 1.07 (0.44–1.91) |
| Maine | 12,894 (6,080–23,484) | 1.18 (0.56–2.15) |
| Massachusetts | 61,890 (37,791–97,579) | 1.12 (0.68–1.77) |
| New Hampshire | 12,640 (7,419–20,782) | 1.15 (0.67–1.89) |
| New Jersey | 77,162 (41,410–120,811) | 1.11 (0.60–1.74) |
| New York | 217,967 (127,370–388,681) | 1.41 (0.83–2.52) |
| Pennsylvania | 123,973 (79,279–194,506) | 1.22 (0.78–1.92) |
| Rhode Island | 13,319 (6,413–24,378) | 1.56 (0.75–2.86) |
| Vermont | 6,335 (3,744–10,739) | 1.24 (0.74–2.11) |
| **South** |  |  |
| Alabama | 44,567 (25,851–70,126) | 1.17 (0.68–1.84) |
| Arkansas | 27,759 (12,686–46,914) | 1.20 (0.55–2.03) |
| Delaware | 11,242 (4,912–20,822) | 1.46 (0.64–2.71) |
| District of Columbia | 12,318 (5,170–22,340) | 2.13 (0.89–3.86) |
| Florida | 266,803 (167,247–545,365) | 1.56 (0.98–3.18) |
| Georgia | 91,816 (42,228–165,513) | 1.14 (0.52–2.05) |
| Kentucky | 49,576 (25,247–80,687) | 1.43 (0.73–2.33) |
| Louisiana | 64,180 (34,109–103,135) | 1.80 (0.96–2.89) |
| Maryland | 59,243 (32,493–107,434) | 1.26 (0.69–2.28) |
| Mississippi | 25,839 (11,874–46,867) | 1.14 (0.52–2.06) |
| North Carolina | 89,965 (53,402–141,224) | 1.11 (0.66–1.74) |
| Oklahoma | 66,621 (29,437–111,712) | 2.22 (0.98–3.72) |
| South Carolina | 45,581 (19,727–75,736) | 1.14 (0.49–1.89) |
| Tennessee | 94,289 (54,997–147,674) | 1.78 (1.04–2.79) |
| Texas | 282,918 (138,179–498,294) | 1.32 (0.65–2.33) |
| Virginia | 76,431 (44,121–119,309) | 1.15 (0.66–1.79) |
| West Virginia | 26,092 (14,731–48,785) | 1.81 (1.02–3.39) |
| **West** |  |  |
| Arizona | 79,527 (45,364–164,022) | 1.42 (0.81–2.93) |
| California | 441,186 (265,134–641,503) | 1.45 (0.87–2.10) |
| Colorado | 58,919 (32,128–95,873) | 1.32 (0.72–2.15) |
| Idaho | 18,367 (10,295–31,404) | 1.39 (0.78–2.37) |
| Montana | 10,740 (5,036–22,036) | 1.28 (0.60–2.63) |
| Nevada | 28,737 (16,677–48,223) | 1.21 (0.70–2.04) |
| New Mexico | 26,094 (14,046–43,214) | 1.61 (0.87–2.67) |
| Oregon | 58,421 (29,106–94,692) | 1.75 (0.87–2.84) |
| Utah | 23,433 (12,742–42,532) | 1.04 (0.57–1.89) |
| Washington | 77,210 (36,291–130,158) | 1.31 (0.61–2.20) |
| Wyoming | 6,524 (3,066–11,051) | 1.46 (0.69–2.48) |

Abbreviations: CrI, credible interval; HCV, hepatitis C virus; DAA: direct-acting antiviral
Note: Estimates represent cumulative period prevalence among adults aged $\geq 18$ years during 2017–2020. Missing outcome values occurred in Rhode Island (n=3); Arizona and the District of Columbia (n=2); and Indiana, North Carolina, Texas, Kansas, and Tennessee (n=1). All missing outcome values were treated as missing and imputed during the MCMC sampling process

**Figure 1.**
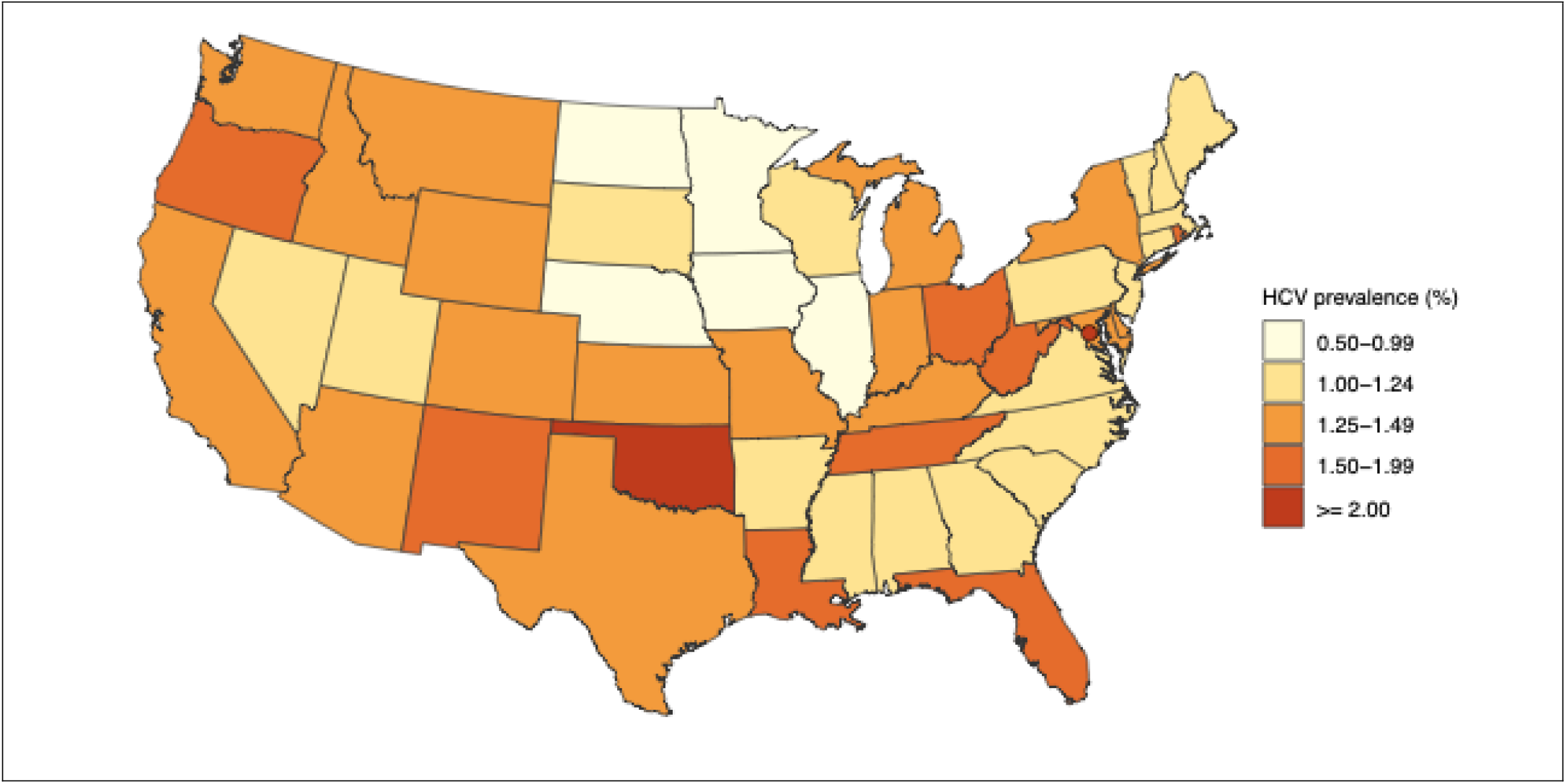
Estimated HCV prevalence (per 100 population) by state, 48 states and the District of Columbia, 2017–2020. Note: Estimated average HCV prevalence across jurisdictions was 1.32%

### Sensitivity analyses

Reporting completeness for surveillance outcomes varied across states: 3 states did not report acute HCV data and 5 contributed data in only one or two years during 2017–2020. Similarly, 6 states did not report chronic HCV data and 7 reported data in only one or two years (eAppendix S1). In the surveillance-excluded analysis, which excluded both acute and chronic HCV surveillance cases, the average state-level prevalence estimate was 1.41% (95% CrI: 0.63%–1.84%). Compared with the main analysis, state-level differences ranged from −0.27 to 1.23 percentage points; only the District of Columbia and Maryland showed increases greater than 0.5 percentage points. Geographic patterns were generally similar between the two models (eTables 1–2, Figure 2).

**Figure 2.**
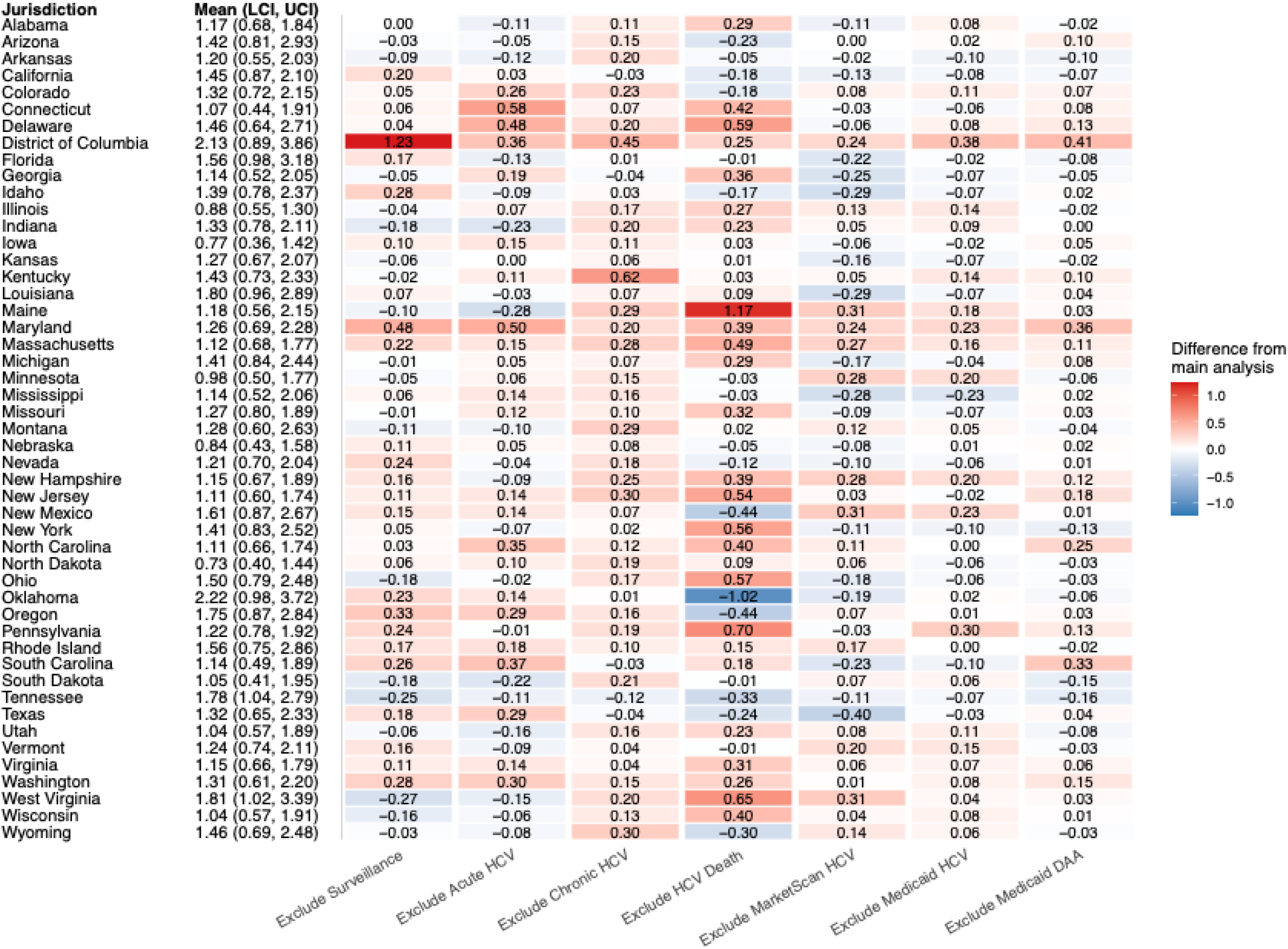
State-level change in estimated HCV prevalence (per 100 population) after excluding individual outcomes, relative to the main analysis. Each cell shows the percentage-point difference in estimated HCV prevalence between an outcome-excluded model and the main analysis: “Exclude Surveillance” corresponds to the surveillance-excluded model, which excluded both acute and chronic HCV surveillance cases, and the subsequent columns correspond to the six leave-one-outcome-out models. Estimates shown in the leftmost column are estimated prevalence from the main analysis (mean, 95% credible interval). Red indicates higher estimated prevalence after excluding the outcome, and blue indicates lower estimated prevalence.

In the leave-one-outcome-out analyses, average prevalence estimates were generally stable when each outcome was removed from the main model. Compared with the main model average of 1.32%, excluding HCV-related deaths produced the largest shift, increasing the average estimate by 0.14 percentage points (eTables 1–2). State-level changes varied by excluded outcome, suggesting that data sources contributed differently across states (Figure 2, eTables 1–2). For example, excluding HCV-related deaths increased estimated prevalence by more than 0.5 percentage points in several states, including Maine, Pennsylvania, West Virginia, Delaware, Ohio, and New Jersey, while reducing the estimate by 1.02 percentage points in Oklahoma.

## Discussion

Using an integrated abundance model and combining hepatitis C–related outcomes derived from multiple data sources, we generated state-level estimates of adult HCV RNA prevalence during 2017– 2020. Our estimates identified geographic variation in HCV burden, with higher estimated state-specific prevalences in some South Central states, Appalachia, and some states in the West Census Region.

These patterns were consistent in a sensitivity analysis excluding surveillance outcome data that resulted in slightly higher estimates overall. Nearly all state-specific prevalence estimates had credible intervals that included the model-averaged national estimate. Therefore, these estimates are well suited for identifying geographic patterns and plausible state-level ranges of HCV burden. Although many of the geographic patterns we observed were similar to the patterns of estimated HCV prevalence from 2013 – 2016, results from the two time periods should not be directly compared due to differences in modeling approaches.^1^□

Standardized state-level estimates of HCV burden are essential data for monitoring elimination efforts. Cost-efficient federal resource allocation requires sub-national estimates to identify geographic areas with the greatest needs for prevention and treatment services. To develop comparable state-level estimates for these purposes, estimates must be produced using similar methods. State-level estimates also help public health practitioners to advocate for policies and resources needed to implement HCV prevention and treatment interventions. This is particularly important given that injection drug use is now the primary risk factor for new HCV infections in the U.S.^36^. People who inject drugs have distinct needs for HCV prevention, including harm reduction services, access to medications for opioid use disorder, and minimally restrictive treatment policies. Policies driving availability of these services are, at least in part, determined at state levels^42^. Estimates of HCV burden can help policy makers understand constituents’ health needs and potentially motivate appropriate state-level investments in elimination.

These estimates also provide a baseline against which states can monitor post-2020 progress toward HCV elimination. However, using them for monitoring will also require on-going burden estimation with a comparable modeling framework. Surveillance data are the only inputs to the model that are produced directly by the jurisdictions, and a higher degree of standardized quality is needed for these data to optimally inform estimates. States and cities that receive funding from CDC for viral hepatitis surveillance have substantially increased their surveillance capacity since funding was expanded in 2021^12^. The percentage of jurisdictions with at least one full-time employee dedicated to hepatitis surveillance increased from 68% to 93% from 2021 to 2025 (N=59). As of July, 2025, however, only 24% of jurisdictions had produced a current HCV prevalence estimate for their jurisdiction, and those were derived from a wide range of methods^12^. Additional, sustained funding is needed for states to routinely produce data to inform standardized HCV burden estimates. The national HIV surveillance system has been producing state-level burden estimates since 2008; these estimates are critical, but they depend on ongoing federal investments. In 2025, federal funding for HIV prevention and treatment was more than $1 billion, compared to $43 million for viral hepatitis^12^.

Most states with higher-than-median HCV point prevalence estimates also had high rates of overdose mortality during this period. Louisiana, New Mexico, and the District of Columbia, which had prevalence estimates more than 20% above the median, also had among the highest jurisdiction-specific overdose mortality rates in the U.S. in 2020^43,44^. West Virginia and Ohio also had prevalence estimates more than 20% above the median and had persistently high overdose mortality rates from 2017 – 2020. Oklahoma, which had the highest prevalence, did not have higher than average overdose mortality during this period, but ranked eighth in the U.S. for opioid dispensing^45^. Oregon had not yet begun experiencing high overdose mortality during 2017-2020, which largely preceded the proliferation of fentanyl in the northwest United States. However, prevalent injection drug use might have contributed to HCV risk during this time.

Our approach integrated multiple aggregate data sources to estimate state-level adult HCV prevalence without requiring individual-level linkage across data sources. We incorporated the previously published national HCV prevalence estimate to help identify the model intercept and anchor the absolute scale of state-level prevalence^1^. Overall, our primary results were robust in leave-one-out sensitivity analyses. The largest differences were observed when HCV-related deaths were excluded, suggesting that mortality data contributed information not fully captured by other HCV-related outcomes. This finding underscores a key advantage of the integrated approach, in which different HCV-related outcomes can contribute complementary information about the population with current infection.

Our results should be interpreted with consideration of several limitations. First, outcomes defined by DAA treatment receipt or HCV-related death indicate current infection during the study period; however, they also reflect individuals leaving the HCV RNA+ population. Our modeling approach used these data sources as signals of state-level variability, and averaged prevalence over four years; this approach might be less sensitive to out- or in-migration changes, as people are cured, die, or newly acquire infections. Second, one of the key model assumptions was that each observed outcome represented a partial count of the underlying population with current HCV infection. However, we cannot rule out the possibility that some outcomes may include false-positive or previous infections, including claims-based coding errors or treatment records misaligned with current infection status.

Third, although we identified and included multiple state-level covariates to account for variation in outcome-specific capture processes and underlying HCV risk factors, some unmeasured structural factors might have contributed to geographic differences in HCV prevalence. For example, federal surveillance funding and years of available surveillance data were included as proxies for surveillance capacity, but variations in surveillance infrastructure and reporting across jurisdictions might not have been fully captured. Adjustment for MarketScan enrollment also might not fully account for state-level differences in coverage or representativeness. Many HCV infections remain undiagnosed; a recent U.S. study estimated that approximately 40% of people with HCV infection were unaware of their infection status^14^. Although we accounted for available state-level sociodemographic characteristics and risk-related covariates to account for these, geographic variation in undiagnosed infection may not have been fully captured because state-level HCV screening practices were not directly observed.

Finally, the national HCV prevalence estimate used to anchor the model included 50 states and the District of Columbia, whereas our analytic sample excluded Alaska and Hawaii to align with the spatial modeling assumptions. Although this difference introduces a minor difference between the population represented by the national anchor and the analytic sample, presented estimates remain comparable across included states and are thus informative.

## Conclusion

We quantified state-level HCV prevalence during 2017–2020. Our estimates should not be interpreted as current prevalence, particularly given recent changes in testing, treatment access, injection drug use–associated risk, and viral hepatitis surveillance capacity. Data reflecting this timeframe serve as an important baseline before expansion of federal funding for viral hepatitis surveillance from 14 to 59 jurisdictions in 2021. Prevalence of HCV infection continues to vary across the United States, with highest burden observed in states that have disproportionately experienced impacts of the opioid crisis and associated increased injection drug use. Elimination of hepatitis C remains an achievable goal, but progress will require investment in prevention, diagnosis and treatment resources, as well as in surveillance systems that provide data for monitoring progress. Composition and magnitude of needs will differ by jurisdiction.

## Data sharing

Data inputs from Merative™ MarketScan® and Medicaid cannot be shared by the authors under the data use agreements. These data are available by inquiry to the respective data holders. The remaining data sources are publicly available and can be accessed through the links provided in the eAppendix.

## Acknowledgement

We thank Dr. Nathan W. Furukawa for sharing aggregated data on direct-acting antiviral use in Medicaid.

