## Supplementary materials for "Estimating Hepatitis C Virus Prevalence in US States and the District of Columbia, 2017–2020"

**eAppendix**

eTable 1. Estimated HCV infection prevalence by state from sensitivity analyses excluding individual and surveillance-based outcomes, 48 states and the District of Columbia, 2017–2020

|  | Surveillance-excluded model | Leave-one-outcome-out models | | | | | |
| --- | --- | --- | --- | --- | --- | --- | --- |
|  | Excluding surveillance cases | Excluding acute HCV surveillance cases | Excluding chronic HCV surveillance cases | Excluding HCV-related death | Excluding MarketScan HCV cases | Excluding Medicaid new HCV diagnoses | Excluding Medicaid DAA treatment receipt |
| Jurisdiction |  | Mean (95% CrI) | | | | | |
| **All** | **1.41 (0.63–1.84)** | **1.40 (0.82–1.99)** | **1.41 (0.89–1.85)** | **1.46 (0.97–1.95)** | **1.25 (0.85–1.78)** | **1.34 (1.04–1.68)** | **1.35 (0.90–1.99)** |
| **Midwest** | | | | | | | |
| Illinois | 0.84 (0.30–1.66) | 0.95 (0.50–2.21) | 1.05 (0.54–2.43) | 1.15 (0.61–2.15) | 1.01 (0.40–2.08) | 1.02 (0.52–1.88) | 0.86 (0.53–1.39) |
| Indiana | 1.15 (0.53–1.78) | 1.10 (0.59–1.93) | 1.53 (0.83–2.57) | 1.56 (0.89–2.65) | 1.38 (0.75–2.70) | 1.42 (0.85–2.46) | 1.33 (0.73–2.35) |
| Iowa | 0.87 (0.38–1.53) | 0.92 (0.40–1.59) | 0.88 (0.54–1.46) | 0.80 (0.44–1.38) | 0.71 (0.40–1.14) | 0.75 (0.45–1.27) | 0.82 (0.44–1.51) |
| Kansas | 1.21 (0.73–1.94) | 1.27 (0.64–1.91) | 1.33 (0.72–2.05) | 1.28 (0.57–2.62) | 1.11 (0.63–1.82) | 1.20 (0.70–1.91) | 1.25 (0.69–1.89) |
| Michigan | 1.40 (0.52–2.42) | 1.46 (0.71–2.55) | 1.48 (0.67–2.21) | 1.70 (0.85–3.05) | 1.24 (0.61–1.93) | 1.37 (0.87–2.64) | 1.49 (0.81–2.94) |
| Minnesota | 0.93 (0.38–1.47) | 1.04 (0.46–2.13) | 1.13 (0.59–1.78) | 0.95 (0.40–1.54) | 1.26 (0.64–2.33) | 1.18 (0.66–1.87) | 0.92 (0.49–1.53) |
| Missouri | 1.26 (0.59–2.16) | 1.39 (0.84–2.43) | 1.37 (0.55–1.97) | 1.59 (0.77–2.60) | 1.18 (0.62–1.92) | 1.20 (0.51–1.93) | 1.30 (0.73–1.92) |
| Nebraska | 0.95 (0.48–1.43) | 0.89 (0.36–1.45) | 0.92 (0.47–1.41) | 0.79 (0.38–1.37) | 0.76 (0.40–1.28) | 0.85 (0.47–1.47) | 0.86 (0.47–1.36) |
| North Dakota | 0.79 (0.37–1.43) | 0.83 (0.40–1.85) | 0.92 (0.46–1.56) | 0.82 (0.43–1.72) | 0.79 (0.43–1.51) | 0.67 (0.40–1.13) | 0.70 (0.43–1.35) |
| Ohio | 1.32 (0.70–1.82) | 1.48 (0.83–2.35) | 1.67 (0.96–2.50) | 2.07 (0.95–4.56) | 1.32 (0.75–2.37) | 1.44 (0.94–2.28) | 1.47 (0.84–2.27) |
| South Dakota | 0.87 (0.42–1.41) | 0.83 (0.33–1.84) | 1.26 (0.54–2.09) | 1.04 (0.46–2.38) | 1.12 (0.51–2.01) | 1.11 (0.61–1.85) | 0.90 (0.46–1.64) |
| Wisconsin | 0.88 (0.35–1.26) | 0.98 (0.47–2.01) | 1.17 (0.62–1.98) | 1.44 (0.66–2.53) | 1.08 (0.52–1.99) | 1.12 (0.64–1.97) | 1.05 (0.54–1.71) |
| **Northeast** | | | | | | | |
| Connecticut | 1.13 (0.46–2.00) | 1.65 (0.79–3.86) | 1.14 (0.60–2.13) | 1.49 (0.59–3.35) | 1.04 (0.45–1.86) | 1.01 (0.44–1.89) | 1.15 (0.50–1.89) |
| Maine | 1.08 (0.23–2.00) | 0.90 (0.37–1.69) | 1.47 (0.64–2.56) | 2.35 (1.02–4.46) | 1.49 (0.58–3.33) | 1.36 (0.66–3.12) | 1.21 (0.50–2.52) |
| Massachusetts | 1.34 (0.33–2.86) | 1.27 (0.67–2.49) | 1.40 (0.59–2.20) | 1.61 (0.69–4.70) | 1.39 (0.65–2.62) | 1.28 (0.53–2.76) | 1.23 (0.66–2.05) |
| New Hampshire | 1.31 (0.36–2.56) | 1.06 (0.48–1.99) | 1.40 (0.62–2.20) | 1.54 (0.67–3.57) | 1.43 (0.68–2.75) | 1.35 (0.73–2.38) | 1.27 (0.62–2.43) |
| New Jersey | 1.22 (0.41–2.09) | 1.25 (0.66–3.03) | 1.41 (0.72–2.68) | 1.65 (0.82–3.06) | 1.14 (0.62–1.91) | 1.09 (0.51–1.97) | 1.29 (0.73–2.14) |
| New York | 1.46 (0.41–2.47) | 1.34 (0.48–2.42) | 1.43 (0.72–2.31) | 1.97 (1.07–3.96) | 1.30 (0.63–2.18) | 1.31 (0.88–1.93) | 1.28 (0.77–2.00) |
| Pennsylvania | 1.46 (0.44–2.43) | 1.21 (0.69–2.01) | 1.41 (0.81–2.18) | 1.92 (1.14–3.45) | 1.19 (0.69–1.84) | 1.52 (0.89–2.62) | 1.35 (0.84–2.30) |
| Rhode Island | 1.73 (0.58–3.45) | 1.74 (0.68–3.54) | 1.66 (0.78–2.84) | 1.71 (0.52–4.23) | 1.73 (0.81–3.65) | 1.56 (0.65–2.68) | 1.54 (0.78–2.78) |
| Vermont | 1.40 (0.35–2.44) | 1.15 (0.40–2.18) | 1.28 (0.59–2.05) | 1.23 (0.49–2.51) | 1.44 (0.65–2.71) | 1.39 (0.61–2.47) | 1.21 (0.64–2.14) |
| **South** | | | | | | | |
| Alabama | 1.17 (0.54–1.82) | 1.06 (0.52–1.77) | 1.28 (0.67–2.04) | 1.46 (0.81–2.58) | 1.06 (0.52–1.97) | 1.25 (0.66–2.31) | 1.15 (0.66–2.07) |
| Arkansas | 1.11 (0.58–1.84) | 1.08 (0.49–2.03) | 1.40 (0.71–2.24) | 1.15 (0.58–2.04) | 1.18 (0.59–2.06) | 1.10 (0.46–1.69) | 1.10 (0.53–2.04) |
| Delaware | 1.50 (0.56–2.80) | 1.94 (0.86–4.54) | 1.66 (0.80–2.92) | 2.05 (1.03–4.04) | 1.40 (0.75–2.71) | 1.54 (0.77–3.10) | 1.59 (0.83–2.85) |
| District of Columbia | 3.36 (1.23–6.42) | 2.49 (1.00–4.76) | 2.58 (1.19–4.85) | 2.38 (0.78–9.33) | 2.37 (0.82–4.76) | 2.51 (1.09–4.23) | 2.54 (1.28–4.38) |
| Florida | 1.73 (0.60–3.32) | 1.43 (0.78–2.85) | 1.57 (0.86–2.99) | 1.55 (0.81–2.53) | 1.34 (0.76–2.03) | 1.54 (1.12–2.49) | 1.48 (0.85–2.11) |
| Georgia | 1.09 (0.54–2.27) | 1.33 (0.64–2.77) | 1.10 (0.52–1.79) | 1.50 (0.60–3.90) | 0.89 (0.47–1.64) | 1.07 (0.62–2.09) | 1.09 (0.53–1.72) |
| Kentucky | 1.41 (0.92–1.96) | 1.54 (0.70–3.15) | 2.05 (0.92–3.21) | 1.46 (0.74–2.44) | 1.48 (0.61–2.63) | 1.57 (0.76–2.82) | 1.53 (0.64–3.16) |
| Louisiana | 1.87 (1.07–2.92) | 1.77 (0.97–3.02) | 1.87 (0.92–3.10) | 1.89 (0.97–3.98) | 1.51 (0.63–2.70) | 1.73 (0.80–2.84) | 1.84 (0.91–2.80) |
| Maryland | 1.74 (0.62–2.75) | 1.76 (0.82–3.32) | 1.46 (0.79–2.33) | 1.65 (0.75–3.34) | 1.50 (0.75–2.76) | 1.49 (0.75–2.58) | 1.62 (0.89–3.07) |
| Mississippi | 1.20 (0.73–1.96) | 1.28 (0.67–2.71) | 1.30 (0.59–2.02) | 1.11 (0.56–1.94) | 0.86 (0.50–1.51) | 0.91 (0.48–1.44) | 1.16 (0.62–2.25) |
| North Carolina | 1.14 (0.58–1.72) | 1.46 (0.74–2.71) | 1.23 (0.68–1.89) | 1.51 (0.76–2.71) | 1.22 (0.62–2.53) | 1.11 (0.68–1.88) | 1.36 (0.75–2.78) |
| Oklahoma | 2.45 (1.38–4.22) | 2.36 (1.26–3.49) | 2.23 (1.23–3.27) | 1.20 (0.63–1.93) | 2.03 (0.99–3.44) | 2.24 (0.78–3.62) | 2.16 (1.16–3.58) |
| South Carolina | 1.40 (0.82–2.11) | 1.51 (0.81–2.72) | 1.11 (0.52–1.84) | 1.32 (0.61–2.24) | 0.91 (0.43–1.51) | 1.04 (0.58–1.58) | 1.47 (0.76–2.42) |
| Tennessee | 1.53 (0.78–2.70) | 1.67 (0.88–2.88) | 1.66 (0.90–2.83) | 1.45 (0.82–2.47) | 1.67 (0.96–2.58) | 1.71 (1.22–2.62) | 1.62 (0.79–2.63) |
| Texas | 1.50 (0.77–2.25) | 1.61 (0.64–2.61) | 1.28 (0.58–2.11) | 1.08 (0.54–1.89) | 0.92 (0.33–1.47) | 1.29 (0.64–1.96) | 1.36 (0.72–2.17) |
| Virginia | 1.26 (0.40–2.34) | 1.29 (0.72–2.70) | 1.19 (0.77–1.82) | 1.46 (0.72–2.56) | 1.21 (0.55–2.36) | 1.22 (0.74–2.02) | 1.21 (0.68–1.96) |
| West Virginia | 1.54 (0.58–2.64) | 1.66 (1.03–3.38) | 2.01 (0.93–3.40) | 2.46 (1.05–4.58) | 2.12 (1.07–4.34) | 1.85 (1.10–3.15) | 1.84 (1.17–3.11) |
| **West** | | | | | | | |
| Arizona | 1.39 (0.53–2.49) | 1.37 (0.59–2.34) | 1.57 (0.61–2.61) | 1.19 (0.57–2.12) | 1.42 (0.69–2.65) | 1.44 (0.85–2.18) | 1.52 (0.65–2.45) |
| California | 1.65 (0.70–3.04) | 1.48 (0.68–2.33) | 1.42 (0.75–2.57) | 1.27 (0.69–2.25) | 1.32 (0.79–2.08) | 1.37 (0.89–1.97) | 1.38 (0.85–2.08) |
| Colorado | 1.37 (0.85–2.24) | 1.58 (0.82–2.86) | 1.55 (0.98–2.33) | 1.14 (0.53–2.41) | 1.40 (0.69–2.16) | 1.43 (0.84–2.11) | 1.39 (0.71–2.01) |
| Idaho | 1.67 (0.80–2.61) | 1.30 (0.65–2.13) | 1.42 (0.71–2.21) | 1.22 (0.60–2.59) | 1.10 (0.50–1.79) | 1.32 (0.64–2.04) | 1.41 (0.82–2.32) |
| Montana | 1.17 (0.64–1.83) | 1.18 (0.60–2.51) | 1.57 (0.65–2.55) | 1.30 (0.63–2.40) | 1.40 (0.64–2.59) | 1.33 (0.56–2.07) | 1.24 (0.72–2.15) |
| Nevada | 1.45 (0.61–2.59) | 1.17 (0.46–2.00) | 1.39 (0.69–2.30) | 1.09 (0.55–2.10) | 1.11 (0.54–1.91) | 1.15 (0.65–1.89) | 1.22 (0.71–2.24) |
| New Mexico | 1.76 (0.67–2.88) | 1.75 (0.60–3.26) | 1.68 (0.69–2.57) | 1.17 (0.54–1.87) | 1.92 (0.97–3.51) | 1.84 (1.09–2.92) | 1.62 (0.91–2.83) |
| Oregon | 2.08 (1.06–3.42) | 2.04 (0.95–3.24) | 1.91 (0.94–3.32) | 1.31 (0.56–2.62) | 1.82 (0.80–2.81) | 1.76 (0.78–2.87) | 1.78 (1.00–2.67) |
| Utah | 0.98 (0.28–1.52) | 0.88 (0.45–1.58) | 1.20 (0.62–1.87) | 1.27 (0.65–2.37) | 1.12 (0.59–2.02) | 1.15 (0.72–1.73) | 0.96 (0.57–1.48) |
| Washington | 1.59 (0.78–2.55) | 1.61 (0.90–3.13) | 1.46 (0.67–2.36) | 1.57 (0.64–2.86) | 1.32 (0.68–2.14) | 1.39 (0.77–2.54) | 1.46 (0.80–2.61) |
| Wyoming | 1.43 (0.85–2.24) | 1.38 (0.67–2.45) | 1.76 (0.79–2.78) | 1.16 (0.53–2.22) | 1.60 (0.73–2.68) | 1.52 (0.60–2.52) | 1.43 (0.73–2.48) |

Abbreviations: CrI, credible interval; HCV, hepatitis C virus; DAA: direct-acting antiviral

Note: Estimates represent cumulative period prevalence among adults aged ≥18 years during 2017–2020, from models excluding the outcome(s) indicated in each column heading. Values are posterior means with 95% credible intervals.

eTable 2. Estimated number of adults with HCV infection by state and excluded outcome, 48 states and the District of Columbia, 2017–2020

|  | Surveillance-excluded model | Leave-one-outcome-out models | | | | | |
| --- | --- | --- | --- | --- | --- | --- | --- |
|  | Excluding surveillance cases | Excluding acute HCV surveillance cases | Excluding chronic HCV surveillance cases | Excluding HCV-related death | Excluding MarketScan HCV cases | Excluding Medicaid new HCV diagnoses | Excluding Medicaid DAA treatment receipt |
| Jurisdiction | Mean (95% CrI) | | | | | | |
| **All** | **3,560,827**  **(1,591,008,–4,646,753)** | **3,535,573**  **(2,070,836–5,025,564)** | **3,560,827**  **(2,247,614–4,672,007)** | **3,687,097**  **(2,449,647–4,924,548)** | **3,156,761**  **(2,146,598–4,495,228)** | **3,384,048**  **(2,626,426–4,242,687)** | **3,409,302**  **(2,272,868–4,773,023)** |
| **Midwest** | | | | | | | |
| Illinois | 82,588  (29,710–163,156) | 93,439  (48,924–217,459) | 103,460  (52,961–239,490) | 112,817  (59,755–212,167) | 99,212  (39,675–204,598) | 100,153  (51,520–185,225) | 84,675  (52,056–137,019) |
| Indiana | 59,020  (27,187–91,489) | 56,499  (30,322–99,385) | 78,899  (42,772–132,142) | 79,958  (45,837–136,304) | 70,816  (38,354–138,562) | 72,802  (43,690–126,307) | 68,303  (37,718–120,632) |
| Iowa | 21,125  (9,126–36,997) | 22,358  (9,646–38,509) | 21,301  (12,977–35,400) | 19,467  (10,601–33,444) | 17,139  (9,653–27,670) | 18,219  (10,816–30,671) | 19,769  (10,789–36,565) |
| Kansas | 26,747  (16,205–42,848) | 28,148  (14,139–42,231) | 29,375  (15,880–45,269) | 28,371  (12,522–57,969) | 24,516  (13,864–40,275) | 26,523  (15,360–42,218) | 27,665  (15,210–41,808) |
| Michigan | 109,169  (40,718–189,550) | 114,009  (55,589–199,507) | 115,489  (52,783–173,212) | 133,078  (66,286–238,225) | 96,868  (47,952–150,744) | 107,073  (68,357–206,325) | 116,906  (63,601–229,967) |
| Minnesota | 39,975  (16,545–63,505) | 45,006  (19,903–91,860) | 48,850  (25,613–76,738) | 40,912  (17,275–66,628) | 54,465  (27,656–100,396) | 50,787  (28,534–80,840) | 39,583  (21,226–65,880) |
| Missouri | 60,061  (27,867–102,719) | 66,050  (39,912–115,796) | 64,916  (25,971–93,800) | 75,801  (36,707–123,561) | 55,890  (29,431–91,193) | 57,143  (24,220–91,621) | 61,962  (34,609–91,413) |
| Nebraska | 13,813  (6,971–20,684) | 12,864  (5,272–21,027) | 13,307  (6,812–20,491) | 11,414  (5,495–19,880) | 10,990  (5,761–18,520) | 12,313  (6,888–21,319) | 12,421  (6,861–19,734) |
| North Dakota | 4,572  (2,170–8,306) | 4,803  (2,341–10,763) | 5,353  (2,662–9,077) | 4,763  (2,496–10,026) | 4,612  (2,521–8,758) | 3,883  (2,334–6,564) | 4,094  (2,488–7,848) |
| Ohio | 120,282  (63,317–165,965) | 134,799  (75,079–214,099) | 152,202  (87,594–227,482) | 188,281  (86,556–414,434) | 120,224  (68,548–215,951) | 131,223  (85,696–207,519) | 133,551  (76,796–206,226) |
| South Dakota | 5,794  (2,827–9,359) | 5,548  (2,170–12,242) | 8,405  (3,571–13,909) | 6,914  (3,042–15,843) | 7,432  (3,421–13,364) | 7,385  (4,034–12,321) | 5,978  (3,073–10,913) |
| Wisconsin | 40,020  (15,800–57,182) | 44,428  (21,404–91,199) | 53,355  (28,263–89,780) | 65,246  (29,987–114,949) | 48,947  (23,721–90,273) | 50,736  (28,894–89,292) | 47,487  (24,682–77,708) |
| **Northeast** | | | | | | | |
| Connecticut | 31,974  (13,071–56,643) | 30,329  (12,406–54,163) | 31,974  (13,071–56,643) | 46,942  (22,356–109,463) | 32,337  (17,032–60,342) | 42,178  (16,705–94,972) | 29,583  (12,671–52,740) |
| Maine | 11,818  (2,500–21,809) | 12,894  (6,080–23,484) | 11,818  (2,500–21,809) | 9,829  (4,032–18,520) | 16,019  (7,023–27,966) | 25,732  (11,197–48,765) | 16,308  (6,314–36,386) |
| Massachusetts | 74,145  (17,973–158,206) | 61,890  (37,791–97,579) | 74,145  (17,973–158,206) | 69,973  (37,009–137,885) | 77,521  (32,601–121,679) | 88,695  (37,968–259,432) | 76,783  (36,005–144,528) |
| New Hampshire | 14,378  (4,009–28,156) | 12,640  (7,419–20,782) | 14,378  (4,009–28,156) | 11,660  (5,234–21,980) | 15,388  (6,801–24,265) | 16,960  (7,354–39,296) | 15,771  (7,498–30,261) |
| New Jersey | 84,772  (28,705–144,768) | 77,162  (41,410–120,811) | 84,772  (28,705–144,768) | 86,943  (45,746–210,559) | 97,769  (50,038–186,168) | 114,633  (57,195–212,009) | 79,092  (43,133–132,576) |
| New York | 224,551  (63,477–381,538) | 217,967  (127,370–388,681) | 224,551  (63,477–381,538) | 206,591  (73,964–373,318) | 219,981  (110,749–356,419) | 303,918  (165,793–611,542) | 199,943  (96,863–336,690) |
| Pennsylvania | 148,270  (44,510–246,405) | 123,973  (79,279–194,506) | 148,270  (44,510–246,405) | 122,554  (69,580–204,472) | 142,742  (81,922–221,350) | 194,748  (116,233–350,316) | 121,223  (70,494–186,712) |
| Rhode Island | 14,749  (4,974–29,409) | 13,319  (6,413–24,378) | 14,749  (4,974–29,409) | 14,803  (5,818–30,228) | 14,146  (6,630–24,177) | 14,551  (4,465–36,119) | 14,788  (6,898–31,144) |
| Vermont | 7,117  (1,762–12,410) | 6,335  (3,744–10,739) | 7,117  (1,762–12,410) | 5,873  (2,023–11,096) | 6,497  (2,996–10,432) | 6,239  (2,478–12,788) | 7,317  (3,313–13,829) |
| **South** | | | | | | | |
| Alabama | 44,677  (20,463–69,246) | 44,567  (25,851–70,126) | 44,677  (20,463–69,246) | 40,519  (19,879–67,340) | 48,578  (25,470–77,655) | 55,429  (30,909–98,228) | 40,452  (19,710–75,023) |
| Arkansas | 25,755  (13,388–42,648) | 27,759  (12,686–46,914) | 25,755  (13,388–42,648) | 24,900  (11,386–47,004) | 32,325  (16,400–51,830) | 26,681  (13,399–47,121) | 27,403  (13,772–47,628) |
| Delaware | 11,534  (4,292–21,503) | 11,242  (4,912–20,822) | 11,534  (4,292–21,503) | 14,862  (6,604–34,908) | 12,744  (6,132–22,464) | 15,724  (7,878–31,039) | 10,781  (5,766–20,824) |
| District of Columbia | 19,425  (7,114–37,154) | 12,318  (5,170–22,340) | 19,425  (7,114–37,154) | 14,426  (5,806–27,532) | 14,951  (6,851–28,090) | 13,783  (4,494–53,987) | 13,732  (4,742–27,543) |
| Florida | 295,864  (102,926–568,458) | 266,803  (167,247–545,365) | 295,864  (102,926–568,458) | 245,524  (134,438–488,984) | 269,622  (146,601–512,133) | 265,165  (138,388–432,795) | 228,983  (130,126–348,523) |
| Georgia | 87,595  (43,702–182,664) | 91,816  (42,228–165,513) | 87,595  (43,702–182,664) | 107,387  (51,235–223,183) | 88,861  (42,079–144,024) | 120,716  (48,096–314,398) | 72,146  (37,795–132,139) |
| Kentucky | 48,654  (31,829–67,845) | 49,576  (25,247–80,687) | 48,654  (31,829–67,845) | 53,388  (24,342–109,001) | 70,817  (31,763–111,220) | 50,442  (25,552–84,342) | 51,362  (20,940–91,199) |
| Louisiana | 66,702  (38,123–104,215) | 64,180  (34,109–103,135) | 66,702  (38,123–104,215) | 63,124  (34,781–107,495) | 66,604  (32,823–110,480) | 67,520  (34,739–141,872) | 53,910  (22,559–96,311) |
| Maryland | 82,051  (29,086–129,531) | 59,243  (32,493–107,434) | 82,051  (29,086–129,531) | 82,633  (38,796–156,381) | 68,796  (37,086–109,696) | 77,760  (35,095–157,248) | 70,732  (35,328–129,693) |
| Mississippi | 27,227  (16,643–44,535) | 25,839  (11,874–46,867) | 27,227  (16,643–44,535) | 29,045  (15,335–61,734) | 29,583  (13,349–45,937) | 25,167  (12,780–44,204) | 19,591  (11,434–34,437) |
| North Carolina | 92,888  (47,295–139,948) | 89,965  (53,402–141,224) | 92,888  (47,295–139,948) | 118,999  (60,000–220,586) | 100,229  (55,228–153,499) | 122,599  (61,667–220,365) | 99,109  (50,794–206,190) |
| Oklahoma | 73,419  (41,337–126,465) | 66,621  (29,437–111,712) | 73,419  (41,337–126,465) | 70,647  (37,755–104,722) | 66,804  (36,890–98,122) | 36,057  (18,971–57,993) | 60,773  (29,710–103,164) |
| South Carolina | 56,286  (32,867–84,736) | 45,581  (19,727–75,736) | 56,286  (32,867–84,736) | 60,404  (32,565–109,338) | 44,439  (21,035–73,852) | 53,050  (24,565–89,797) | 36,519  (17,210–60,573) |
| Tennessee | 80,864  (41,373–142,907) | 94,289  (54,997–147,674) | 80,864  (41,373–142,907) | 88,194  (46,534–152,256) | 87,833  (47,408–149,644) | 76,493  (43,470–130,797) | 88,265  (50,936–136,471) |
| Texas | 321,274  (165,159–481,486) | 282,918  (138,179–498,294) | 321,274  (165,159–481,486) | 344,810  (136,825–559,548) | 274,304  (123,271–451,346) | 232,352  (115,295–403,869) | 196,817  (69,960–315,164) |
| Virginia | 84,049  (26,757–156,071) | 76,431  (44,121–119,309) | 84,049  (26,757–156,071) | 85,742  (47,737–180,030) | 79,574  (51,538–121,137) | 97,114  (47,935–170,526) | 80,769  (36,900–157,570) |
| West Virginia | 22,145  (8,389–37,977) | 26,092  (14,731–48,785) | 22,145  (8,389–37,977) | 23,912  (14,823–48,590) | 28,909  (13,371–48,908) | 35,411  (15,141–65,906) | 30,524  (15,391–62,356) |
| **West** | | | | | | | |
| Arizona | 77,761  (29,828–138,997) | 79,527  (45,364–164,022) | 77,761  (29,828–138,997) | 76,522  (32,860–130,543) | 88,007  (33,875–145,733) | 66,642  (32,010–118,448) | 79,626  (38,308–148,047) |
| California | 503,638  (214,871–924,657) | 441,186  (265,134–641,503) | 503,638  (214,871–924,657) | 450,708  (207,361–711,318) | 431,646  (227,488–782,040) | 385,845  (209,239–685,090) | 402,867  (241,376–635,078) |
| Colorado | 61,029  (38,037–100,027) | 58,919  (32,128–95,873) | 61,029  (38,037–100,027) | 70,430  (36,376–127,648) | 69,127  (43,528–104,042) | 50,722  (23,764–107,426) | 62,640  (30,986–96,445) |
| Idaho | 22,087  (10,565–34,569) | 18,367  (10,295–31,404) | 22,087  (10,565–34,569) | 17,202  (8,645–28,242) | 18,827  (9,377–29,210) | 16,173  (7,900–34,358) | 14,605  (6,568–23,737) |
| Montana | 9,781  (5,363–15,362) | 10,740  (5,036–22,036) | 9,781  (5,363–15,362) | 9,915  (5,037–20,972) | 13,146  (5,445–21,315) | 10,871  (5,314–20,120) | 11,746  (5,377–21,692) |
| Nevada | 34,400  (14,571–61,236) | 28,737  (16,677–48,223) | 34,400  (14,571–61,236) | 27,677  (10,933–47,401) | 32,899  (16,250–54,486) | 25,760  (13,043–49,841) | 26,225  (12,788–45,204) |
| New Mexico | 28,547  (10,851–46,627) | 26,094  (14,046–43,214) | 28,547  (10,851–46,627) | 28,299  (9,639–52,750) | 27,139  (11,113–41,655) | 18,938  (8,671–30,310) | 30,994  (15,684–56,843) |
| Oregon | 69,153  (35,212–113,877) | 58,421  (29,106–94,692) | 69,153  (35,212–113,877) | 67,867  (31,610–107,758) | 63,508  (31,402–110,440) | 43,628  (18,654–87,442) | 60,561  (26,686–93,501) |
| Utah | 22,058  (6,370–34,189) | 23,433  (12,742–42,532) | 22,058  (6,370–34,189) | 19,824  (10,032–35,424) | 26,941  (14,047–41,995) | 28,490  (14,519–53,194) | 25,102  (13,222–45,339) |
| Washington | 94,115  (46,040–150,839) | 77,210  (36,291–130,158) | 94,115  (46,040–150,839) | 95,264  (53,198–184,674) | 86,069  (39,694–139,172) | 92,444  (37,782–168,819) | 78,130  (39,880–126,340) |
| Wyoming | 6,374  (3,795–9,990) | 6,524  (3,066–11,051) | 6,374  (3,795–9,990) | 6,154  (3,003–10,948) | 7,866  (3,517–12,390) | 5,179  (2,366–9,879) | 7,134  (3,264–11,930) |

Abbreviations: CrI, credible interval; HCV, hepatitis C virus; DAA: direct-acting antiviral

Note: Estimates represent the cumulative number of adults aged ≥18 years during 2017–2020, from models excluding the outcome(s) indicated in each column heading. Values are posterior means with 95% credible intervals

**eAppendix S1. Data sources and variables definitions**

**S1.1 HCV-related outcomes definition**

*MarketScan HCV cases*

MarketScan HCV cases were derived from the Merative™ MarketScan® Commercial and Medicare Databases for 2017–2020. HCV cases were defined as individuals aged ≥18 years with at least one inpatient or outpatient claim containing an HCV diagnosis code or at least one pharmacy claim for dispensed DAA treatment during the study period. Geographic assignment was based primarily on the enrollee’s state of residence. When state of residence was unavailable, metropolitan statistical area of residence or inpatient facility state was used. For metropolitan statistical areas spanning more than one state, individuals were assigned to the dominant state. To maximize the availability of geographic information, individuals were counted once, based on their earliest HCV-related claim with available geographic information during the study period. Approximately 10% of cases had missing geographic information and were excluded in a complete-case analysis. HCV diagnosis codes and DAA National Drug Codes used to define cases are listed below.

| Variable | Code |
| --- | --- |
| HCV diagnosis | ICD-10 code:  B17.10; B17.11; B18.2; B19.20; B19.21 |
| DAA treatment | National Drug Codes (11-digit codes):  00006307402, 00074260028, 00074262504, 00074262501, 00074262528, 00074262556, 00074262580, 00074262584, 61958180101, 61958180301, 61958180401, 61958180206, 61958180501, 72626260101, 61958150101, 61958150301, 61958150401, 61958150202, 61958150501, 61958240101, 61958220101, 61958220301, 61958220401, 61958220501, 61958220201, 61958220202, 61958220203, 72626270101, 00003001101, 00003021301, 00003021501, 00074006301, 00074006328, 00074309301, 00074309328, 00074308228, 59676022507, 59676022528, 61958150201, 61958180202, 00006307401, 61958220402, 51167010001, 51167010003, 00085031402. |

*Medicaid new HCV diagnoses*

Medicaid new HCV diagnoses were based on aggregated state-level counts from a prior study (Kapadia SN et al., 2023) using 2017–2019 Transformed Medicaid Statistical Information System Analytic Files. The source study identified adults aged 18–64 years with a new HCV diagnosis using an HCV RNA test followed by an HCV diagnosis code within 180 days, with a 12-month look-back period to exclude prior diagnoses. Individuals were required to be continuously enrolled in Medicaid and not dually enrolled in Medicare; Rhode Island, Tennessee, and Kansas were excluded from the source study because of missing key demographic data.

*Medicaid DAA treatment receipt*

Medicaid DAA treatment receipt was based on a prior study (Furukawa NW et al.,) that used CMS Medicaid Analytic eXtract and Transformed Medicaid Statistical Information System Analytic Files to identify Medicaid beneficiaries who filled prescriptions for HCV direct-acting antivirals. The source study included Medicaid prescription claims across Medicaid-supported plan types and linked beneficiaries longitudinally across data systems using CMS identifiers. Individuals were counted once, in the state and year of their first observed DAA prescription fill; subsequent treatment courses were not counted. State-year counts fewer than 10 were suppressed in the source data. The specific medication codes used to define DAA treatment are available in the source study supplement.

***States and jurisdictions with missing HCV-related outcome values***

| State or jurisdictions | Number of missing outcomes | Missing outcomes |
| --- | --- | --- |
| Rhode Island | 3 | Acute HCV surveillance cases; chronic HCV surveillance cases; Medicaid new HCV diagnoses |
| Arizona, District of Columbia | 2 | Acute HCV surveillance cases; chronic HCV surveillance cases |
| Indiana, North Carolina, Texas | 1 | chronic HCV surveillance cases |
| Kansas, Tennessee | 1 | Medicaid new HCV diagnoses |

**S1.2 Data sources and definitions for covariates**

| Covariate | Data source | Definition/ Note | Model role |
| --- | --- | --- | --- |
| Average annual surveillance funding received | [Tracking Accountability in Government Grants System](https://taggs.hhs.gov/) | Average annual amount of surveillance funding received during 2017-2020. | Outcome-specific covariate: acute HCV and chronic HCV surveillance cases |
| Years with available acute HCV surveillance data | [Viral Hepatitis Surveillance Reports](https://www.cdc.gov/hepatitis/statistics/2021surveillance/data/xlsx/table-3.1.xlsx) | Number of years during 2017-2020 with acute HCV surveillance data reported to CDC (range: 0-4). | Outcome-specific covariate: acute HCV surveillance cases |
| Years with available chronic HCV surveillance data | Viral Hepatitis Surveillance Reports ([2017](https://stacks.cdc.gov/view/cdc/81817), [2018](https://archive.cdc.gov/www_cdc_gov/hepatitis/statistics/2018surveillance/HepC.htm), [2019](https://www.cdc.gov/hepatitis/statistics/2019surveillance/pdfs/2019HepSurveillanceRpt.pdf), [2020](https://www.cdc.gov/hepatitis/statistics/2020surveillance/hepatitis-c/table-3.5.htm)) | Number of years during 2017-2020 with chronic HCV surveillance data reported to CDC (range: 0-4). | Outcome-specific covariate: chronic HCV surveillance cases |
| Population served by a medical examiner, % | Medicolegal death investigation systems ([2018](https://www.cdc.gov/comec/about/index.html.)) | Percentage of the state population served by a medical examiner. County-level 2018 data were weighted by county population and aggregated to the state level; 2018 values were used to represent the study period. | Outcome-specific covariate: HCV-related death |
| State medical examiner oversight |  | Indicator for whether the state had a state medical examiner in addition to county officials (yes=1, no=0). 2018 values were used to represent the study period. | Outcome-specific covariate: HCV-related death |
| MarketScan enrollees relative to adult population | MarketScan enrollment files | Ratio of unique MarketScan enrollees to the adult population during 2017–2020, calculated from MarketScan enrollment files and CDC WONDER population estimates. To align with the MarketScan HCV case definition, enrollees were assigned geographic information based on the earliest enrollment record with available state of residence or metropolitan statistical area. | Outcome-specific covariate: MarketScan HCV cases |
| Age distribution, %  • Ages 18-40 years  • Ages 41-54 years  • Ages 55-64 years  • Ages 65 years and older | American Community Survey 1-year estimates, accessed through the [tidycensus](https://cran.r-project.org/web/packages/tidycensus/index.html) package for 2017–2019; 2020 ACS 1-year experimental data, accessed from the [Census Bureau website](https://www.census.gov/programs-surveys/acs/data/experimental-data.html). | Average annual percentage during 2017-2020, calculated as persons in each age group divided by the total population. | Outcome-specific covariate: MarketScan HCV cases, Medicaid new HCV diagnoses,  Medicaid DAA treatment receipt; Prevalence covariates |
| Unemployment, % |  | Average annual percentage during 2017-2020, calculated as unemployed persons divided by the total labor force. | Outcome-specific covariate: MarketScan HCV cases; Prevalence covariates |
| Population living below the poverty line, % |  | Average annual percentage during 2017-2020, calculated as persons living below the poverty line divided by persons with known poverty status. | Outcome-specific covariate: MarketScan HCV cases; Prevalence covariates |
| Population with at least a high school diploma, % |  | Average annual percentage during 2017-2020, calculated as persons aged 25 years or older with at least a high school diploma divided by the population aged 25 years or older. | Outcome-specific covariate: MarketScan HCV cases;  Prevalence covariates |
| Households receiving public assistance, % |  | Percentage of households receiving public assistance divided by total households. Values were averaged over available years (2017-2019); 2020 values were unavailable. | Outcome-specific covariate: MarketScan HCV cases;  Prevalence covariates |
| Median household income |  | Average annual state-level median household income during 2017-2020. | Outcome-specific covariate: MarketScan HCV cases;  Prevalence covariates |
| Male population, % |  | Average annual percentage during 2017-2020, calculated as males divided by the total population. | Prevalence covariates |
| Black population, % |  | Average annual percentage during 2017-2020, calculated as Black population divided by the total population. | Prevalence covariates |
| Hispanic population, % |  | Average annual percentage during 2017-2020, calculated as Hispanic population divided by the total population. | Prevalence covariates |
| Medicaid enrollment as a percentage of adult population | Furukawa NW, Ingber SZ, Symum H, et al. Medicaid Expansion and Restriction Policies for Hepatitis C Treatment. *JAMA Network Open*. 2024;7(7):e2422406. | Medicaid enrollment divided by the adult population. | Outcome-specific covariate: MarketScan HCV cases, Medicaid new HCV diagnoses,  Medicaid DAA treatment receipt |
| Medicaid expansion status |  | Indicator for whether the state had expanded Medicaid (yes=1, no=0). | Outcome-specific covariate: Medicaid new HCV diagnoses,  Medicaid DAA treatment receipt |
| Years without fibrosis restrictions for DAA prescription |  | Number of years during 2017-2020 without fibrosis restrictions for DAA prescription (range: 0-4). Fibrosis restrictions were categorized as none, requiring F1-F2 fibrosis, or requiring F3-F4 fibrosis. | Outcome-specific covariate: Medicaid new HCV diagnoses,  Medicaid DAA treatment receipt |
| Years without sobriety restrictions for DAA prescription |  | Number of years during 2017-2020 without sobriety restrictions for DAA prescription (range: 0-4). Sobriety restrictions were categorized as none, screening or counseling for substance use disorder, 1-5 months of sobriety, or 6-12 months of sobriety. | Outcome-specific covariate: Medicaid new HCV diagnoses,  Medicaid DAA treatment receipt |
| Years without prescriber restrictions for DAA prescription |  | Number of years during 2017-2020 without prescriber restrictions for DAA prescription (range: 0-4). Prescriber restrictions were categorized as none, specialist consult, or specialist required. | Outcome-specific covariate: Medicaid new HCV diagnoses,  Medicaid DAA treatment receipt |
| Unsheltered homelessness rate per 100,000 persons | [United States Department of Housing and Urban Development](https://www.huduser.gov/portal/datasets/ahar/2024-ahar-part-1-pit-estimates-of-homelessness-in-the-us.html) | Average annual unsheltered homeless population per 100,000 persons during 2017-2020. | Prevalence covariates |
| Adult incarceration rate per 100,000 persons | [Correctional populations in the United States, U.S. Department of Justice, Office of Justice Programs, Bureau of Justice Statistics](https://bjs.ojp.gov/library/publications/correctional-populations-united-states-2017-2018) | Average annual number of incarcerated individuals aged 18 years or older per 100,000 persons during 2017-2020. | Prevalence covariates |
| Drug overdose death rate per 100,000 persons | [National Vital Statistics System](https://www.cdc.gov/nchs/nvss/vsrr/drug-overdose-data.htm) | Average annual drug overdose deaths per 100,000 persons during 2017-2020. Drug overdose deaths were identified using ICD-10 underlying cause-of-death codes X40-X44, X60-X64, X85, and Y10-Y14. | Prevalence covariates |
| Treatment admissions reporting injection drug use per 100,000 persons | [Treatment Episode Data Set-Admissions](https://www.samhsa.gov/data/data-we-collect/teds-treatment-episode-data-set/datafiles/teds-a-2006-2023) | Average annual treatment admissions reporting injection drug use per 100,000 persons during 2017-2020. | Prevalence covariates |

**eAppendix S2. Model Description and implementation**

**S2.1 Bayesian spatial integrated abundance model**

The Bayesian spatial integrated abundance model used in our analysis is highly adapted from the model proposed by Hepler et al^[1]^. Except that we did not consider the temporal effect in our model. The outcomes, hence, are comprised of the total number of individuals over the given years. We present our model in detail below.

Let $Y_{i}^{k}$ denote the total observed count for outcome $k$ $(k=1,\cdots,K)$ in state $i$ $(i=1,\cdots,n)$ across a given time interval $\boldsymbol{t}$, and $N_{i}$ be the latent number of people with HCV infection in state $i$ during $\boldsymbol{t}$. For each outcome $k$, we assume

$$Y_{i}^{k}|N_{i},p_{i}^{k} \sim\mathrm{Binomial}\left( N_{i},p_{i}^{k} \right),$$

(S1)

where $p_{i}^{k}$represents the proportion of people with HCV infection who experience outcome $k$ in state $i$ during $\boldsymbol{t}$. It is modelled using the logistic regression

$\mathrm{logit}\left( p_{i}^{k} \right)=\beta_{0}^{k}+\boldsymbol{X}_{i}^{k}\boldsymbol{\beta}^{k}+f_{i}^{k}+\epsilon_{i}^{k}$,

(S2)

where $\beta_{0}^{k}$ is the intercept, $\boldsymbol{X}_{i}^{k}$ is the covariate vector related to state $i$, $\boldsymbol{\beta}^{k}$ is the coefficient vector corresponding to $\boldsymbol{X}_{i}^{k}$, $f_{i}^{k}$ refers to the spatial random effect, and $\epsilon_{i}^{k}$ is the error term. The spatial random effect $f_{i}^{k}$ is assumed to follow the intrinsic conditional autoregressive (ICAR) model^[4]^, which is

$f_{i}^{k}|\boldsymbol{f}_{-i}^{k},\tau_{k}^{2} \sim N\left( \frac{1}{\omega_{i+}}\sum_{j\neq i} \omega_{ij}f_{j}^{k}, \frac{\tau_{k}^{2}}{\omega_{i+}} \right),$,

(S3)

$$\tau_{k}^{2}\sim\mathrm{IG}\left( a,b \right),$$

where $\boldsymbol{f}_{-i}^{k}=\left\{ f_{j}^{k}:j\neq i \right\}$, $\omega_{ij}$ is a binary indicator that takes the value 1 or 0 to indicate if states $i$ and $j$ share a border or not, $\omega_{i+}=\sum_{j} \omega_{ij}$ is the total number of neighbours for the state $i$, and $\tau_{k}^{2}$ is related to the conditional variance with an Inverse-Gamma (IG) prior. To account for the potential dependencies among the outcomes, the error terms: $\boldsymbol{\epsilon}_{i}=(\epsilon_{i}^{1},\cdots,\epsilon_{i}^{K})$, for each state $i$ across all outcomes $k=1,\cdots,K$, are assumed to be from a multivariate normal distribution with mean zero^[3]^:

$$\boldsymbol{\epsilon}_{i}|\boldsymbol{\Sigma}\sim N\left( \boldsymbol{0},\boldsymbol{\Sigma} \right),$$

$$\boldsymbol{\Sigma\sim}\mathrm{Wishart} \left( \boldsymbol{I}_{K}\mathbf{,}K \right)\boldsymbol{,}$$

where $\boldsymbol{\Sigma}$ refers to the covariance matrix and is assigned a conjugate Wishart prior distribution.

For the latent number of people with HCV infection $(N_{i})$, we assume

$$N_{i}|\lambda_{i}\sim\mathrm{Binomial}\left( \bar{P}_{i},\mu\lambda_{i} \right),$$

where $\bar{P}_{i}$ denotes the averaged total population size in state $i$ across $\boldsymbol{t}$**,** $\mu$ refers to the nationwide prevalence of hepatitis C during $\boldsymbol{t}$, and $\lambda_{i}$ is the prevalence ratio of HCV infection in state $i$ compared with $\mu$. The prevalence ratio $\lambda_{i}$ is modelled using the log-linear regression

$\log\left( \lambda_{i} \right)=\eta_{0}+\boldsymbol{W}_{i}\boldsymbol{\eta}+\theta_{i}+\upsilon_{i}$,

(S4)

where $\eta_{0}$ is the intercept, $\boldsymbol{W}_{i}$ is the covariate vector related to state $i$, $\boldsymbol{\eta}$ is the corresponding coefficient vector, $\theta_{i}$ is the spatial random effect, and

$$\upsilon_{i}|\sigma_{\upsilon}^{2} \sim N(0,\sigma_{\upsilon}^{2})$$

accounts for the unexplained variance. Similarly, we set

$$\sigma_{\upsilon}^{2}\sim\mathrm{IG}\left( a,b \right)$$

as the prior for $\sigma_{\upsilon}^{2}$. The spatial random effects $\theta_{i}$ are also assumed to follow the ICAR model:

$$\theta_{i}|\boldsymbol{\theta}_{-i},\tau_{\theta}^{2} \sim N\left( \frac{1}{\omega_{i+}}\sum_{j\neq i} \omega_{ij}\theta_{j}, \frac{\tau_{\theta}^{2}}{\omega_{i+}} \right),$$

(S5)

$$\tau_{\theta}^{2} \sim\mathrm{IG}\left( a,b \right),$$

where $\boldsymbol{\theta}_{-i}=\left\{ \theta_{j}:j\neq i \right\}$, and $\tau_{\theta}^{2}$ is related to the conditional variance with an Inverse-Gamma (IG) prior. In addition, for all intercepts and the regression coefficient parameters: $\beta_{0}^{k}$, $\boldsymbol{\beta}^{k}$, $\eta_{0}$, and $\boldsymbol{\eta}$ in models (S2) and (S4), we assume flat and improper prior distributions over the real line. In terms of the hyper-parameter setting for the IG priors, we use $a=b=0.5$.

**2.2 The posterior distribution**

Following the above proposed model, the full joint posterior distribution can be written as

$$\prod_{i} \prod_{k} p\left( N_{i},p_{i}^{k},\lambda_{i} | Y_{i}^{k},\bar{P}_{i},\mu\right)=\left\{ \prod_{i} \left[ \prod_{k} p\left( Y_{i}^{k} | N_{i},p_{i}^{k} \right)p\left( \beta_{0}^{k} \right)p\left( \boldsymbol{\beta}^{k} \right)p\left( f_{i}^{k} | \boldsymbol{f}_{-i}^{k},\tau_{k}^{2} \right)p\left( \tau_{k}^{2} \right) \right]\times p\left( \boldsymbol{\epsilon}_{i} | \boldsymbol{\Sigma} \right)p\left( N_{i} | \bar{P}_{i},\mu, \lambda_{i} \right)p\left( \theta_{i} | \boldsymbol{\theta}_{-i},\tau_{\theta}^{2} \right)p\left( \upsilon_{i} | \sigma_{\upsilon}^{2} \right) \right\}p\left( \boldsymbol{\Sigma} \right)p\left( \eta_{0} \right)p\left( \boldsymbol{\eta} \right)p\left( \tau_{\theta}^{2} \right)p\left( \sigma_{\upsilon}^{2} \right).$$

Both the latent number of people with HCV in each state $(N_{i})$ and the state-level prevalence ratio of HCV infection ${(\lambda}_{i})$ can be estimated using the draws sampled from this posterior distribution, given the data input $(Y_{i}^{k},\bar{P}_{i},\mu)$, $i=1,\cdots,n$, and $k=1,\cdots,K$, during the time interval $\boldsymbol{t}$**.**

**2.3 Computation**

An adaptive Metropolis-within-Gibbs Markov chain Monte Carlo (MCMC) algorithm has been implemented for posterior sampling using the *R* package *NIMBLE* ^[5]^. Ten MCMC chains were set up with different random initialisations. For each of the chains, we set the total number of iterations to 100,000 with the first half as burn-in and thinning set to 5. Convergence was checked using trace plots and the modified Gelman–Rubin diagnostic statistics^[6]^. This resulted in 100,000 MCMC draws available to use for the estimations of the latent number of people with HCV $(N_{i})$ and the state-level prevalence of HCV ${(\mu\lambda}_{i})$.

To ensure the posterior distribution is proper, a centering or sum-to-zero constraint was enforced for the ICAR models (S3) and (S5) during the MCMC sampling process. Due to these modelling constraints inherent in the ICAR models, we excluded Alaska and Hawaii from our analyses, as they are geographically separate from the contiguous United States (and thus disjoint from other states). The centering constraint cannot be applied when there are disjoint groups of regions, as each requires a distinct improper flat prior on the mean[5].

The missing values in some outcomes for certain states were imputed based on the model (S1) during the MCMC sampling by the *NIMBLE* package in *R*.
